# Decoding phantom limb movements from intraneural recordings

**DOI:** 10.1101/2025.08.21.25333903

**Authors:** Cecilia Rossi, Marko Bumbasirevic, Paul Čvančara, Thomas Stieglitz, Stanisa Raspopovic, Elisa Donati, Giacomo Valle

## Abstract

Limb loss leads to severe sensorimotor deficits and requires the use of a prosthetic device, especially in lower-limb amputees. While direct recording from residual nerves offers a biomimetic route for an effective prosthetic control, the low amplitude and noisy nature of these neural signals together with the challenge of establishing a reliable nerve interfacing, have hindered its adoption. Intraneural multichannel electrodes could potentially establish an effective interface with the nerve fibers, enabling access to motor signals even from muscles lost after the amputation.

In this study, we report the direct neural recordings of two transfemoral amputees using transversal intrafascicular multichannel electrodes (TIME) implanted in the tibial nerves. We observed multiunit activity associated with volitional phantom movements of the knee, ankle and toes flexion and extension, with joint- and direction-specific neural modulation in both participants. The motor signals were distributed across all the electrodes, showing both single-joint and multi-joint selectivity, as well as direction selectivity for limb flexion and extension.

After characterizing the neural evoked activity, we developed a Spiking Neural Network (SNN)-based decoder that outperform conventional motor decoders in predicting attempted phantom leg movements. Decoding accuracy improved further by including a broader signal bandwidth that captured both intraneural (ENG) and inter-muscular (imEMG) activity. Finally, comparing motor maps (recording) with sensory maps (stimulation) revealed a minimal overlap, suggesting early segregation of motor and sensory fibers within the tibial nerve before the knee bifurcation.

Our findings demonstrate the feasibility to record motor signal and decode lower-limb movements directly from the nerves in amputees using intraneural interfaces. This paves the way for bidirectional, neurally-controlled prosthetic limbs combining natural control with somatosensory feedback through a single implanted interface.

## Introduction

The human sensorimotor system relies on a tight interplay between voluntary motor commands and continuous sensory feedback to generate coordinated, dexterous movements. Limb loss completely disrupts the brain-body communication, leading to severe sensorimotor disabilities and psychological distress. As a result, many amputees remain wheelchair bound, immobile, or only partially integrated in the activities of daily life. Lower-limb amputations (LLA) are more prevalent than upper-limb ones, accounting for 69% of all the individuals living with the loss of a limb^1^. Among these, more than half are major amputation^2^, i.e., above the ankle level. In addition, higher level of amputation is associated with increased disability, characterized by reduced walking speed and higher energy expenditure (i.e., above-knee amputation (AKA) or hip-disarticulation)^3^.

LLA prosthetic devices are commonly used to restore mobility, but their functionality remains limited, particularly in terms of intuitive volitional control and somatosensory feedback integration^4^. Current commercial solutions typically rely on passive mechanisms of action, avoiding direct control by the user^5^. In some cases, surface electromyographic signals (sEMG) could be used to directly control the prosthetic device (i.e., myoelectric prostheses)^6^. However, these solutions lack the specificity and robustness needed for fine-grained, volitional control, particularly when multiple degrees of freedom (DoF) are involved. A major barrier to the efficacy of sEMG decoders is the challenge of accessing the appropriate muscles. Muscles located deep within the thigh, those that have been anatomically reorganized post-amputation, and those lost due to the amputation present significant challenges for capturing reliable movement-related signals through surface recordings^7^.

To address this important limitation of non-invasive techniques, novel surgical approaches involving nerve or muscle transfer have been developed and successfully tested for decoding attempted phantom movements^8^: Regenerative peripheral nerve interfaces (RPNIs)^9^ and Target muscles reinnervation (TMR)^10^. While these approaches have shown promise, they have primarily been evaluated for motor decoding in upper-limb amputees (RPNIs)^9,11^ or a single AKA with a few DoFs^12^. Agonist-antagonist Myoneural Interfaces (AMI) have been successfully used in multiple transtibial amputees, significantly improving their gait^13,14^. However, only two studies have reported AMI in pilot trials with AKA, with promising results^15,16^, though further testing is still necessary to validate the approach. While these approaches have successfully decoded movements of the knee or ankle, none have demonstrated the ability to decode movements from all the leg joints, including the foot and toes.

In addition to motor control, somatosensory feedback from the leg and foot is essential for providing meaningful information to the users’ brain, supporting sensorimotor control^17^, posture, and balance^18^. Recent studies have shown the use of neural interfaces implanted in the tibial, peroneal, and sciatic nerves after limb loss for restoring rich and selective somatosensory information. Results showed multiple functional and cognitive benefits in both transtibial^19,20^ and transfemoral amputees^21–25^, highlighting the importance of restoring sensory feedback alongside motor control. The integration of the somatosensory feedback with the motor decoding system could further enhance the control and usability of prostheses, creating a bidirectional communication system that mimics natural limb function. Although the implantable electrodes are in close contact with, not only afferent, but also efferent fibers encoding information about the volitional limb movement, none of these electrodes has been used to record electroneurographic (ENG) signals for motor decoding. This is primarily because ENG signals are low in amplitude and highly prone to noise, requiring penetrating electrodes, such as Utah arrays^26^ or microneurography needles^27^, to capture them reliably and advanced real-time decoders, which have so far hindered the development of neurally-controlled prostheses. Few attempts only in upper-limb amputees, implanted in the median and ulnar nerves, have shown promising off-line decoding of fingers and hand movements^26,28–30^. To date, no studies have attempted to record neural signals and decode leg movements in humans. The ability to directly record and decode the neural activity from peripheral nerves could provide access to a richer set of control signals for prosthetics, including those associated with muscles lost after amputation, enabling faster and more complex movements.

To this aim, we implanted multiple TIMEs in two individuals with AKA in their tibial branch of the sciatic nerve (**Figure 1A, B**). We asked the participants to move their phantom leg, including knee, ankle and toes, in different directions (**Figure 1C**), while simultaneously recording the intraneural activity (**Figure 1D**). Unlike prior studies where TIMEs were used solely for stimulation, we employ them here as a bidirectional interface, capturing motor intent and delivering sensory feedback through the same intraneural sites in AKA. We used this dataset to conduct a comprehensive analysis of motor decoding and sensory mapping. Firstly, we comprehensively characterized the neural efferent response in different movement conditions (e.g., joint and direction selectivity), including both the neural dynamics and the spatial activation of the 56 implanted active sites within the nerve. Secondly, we designed and implemented a purposely-designed intraneural motor decoder, based on a spiking neural network. The SNN, aligned with the event-driven structure of ENG signals, significantly outperformed standard classifiers in both accuracy and efficiency in both participants^31^. In addition, we also demonstrated the possibility to use this implantable technology to record inter-muscular activity. Finally, we compared motor maps derived from the intraneural recordings with the sensory maps obtained delivering electrical neurostimulation through the same electrodes.

**Figure 1.**
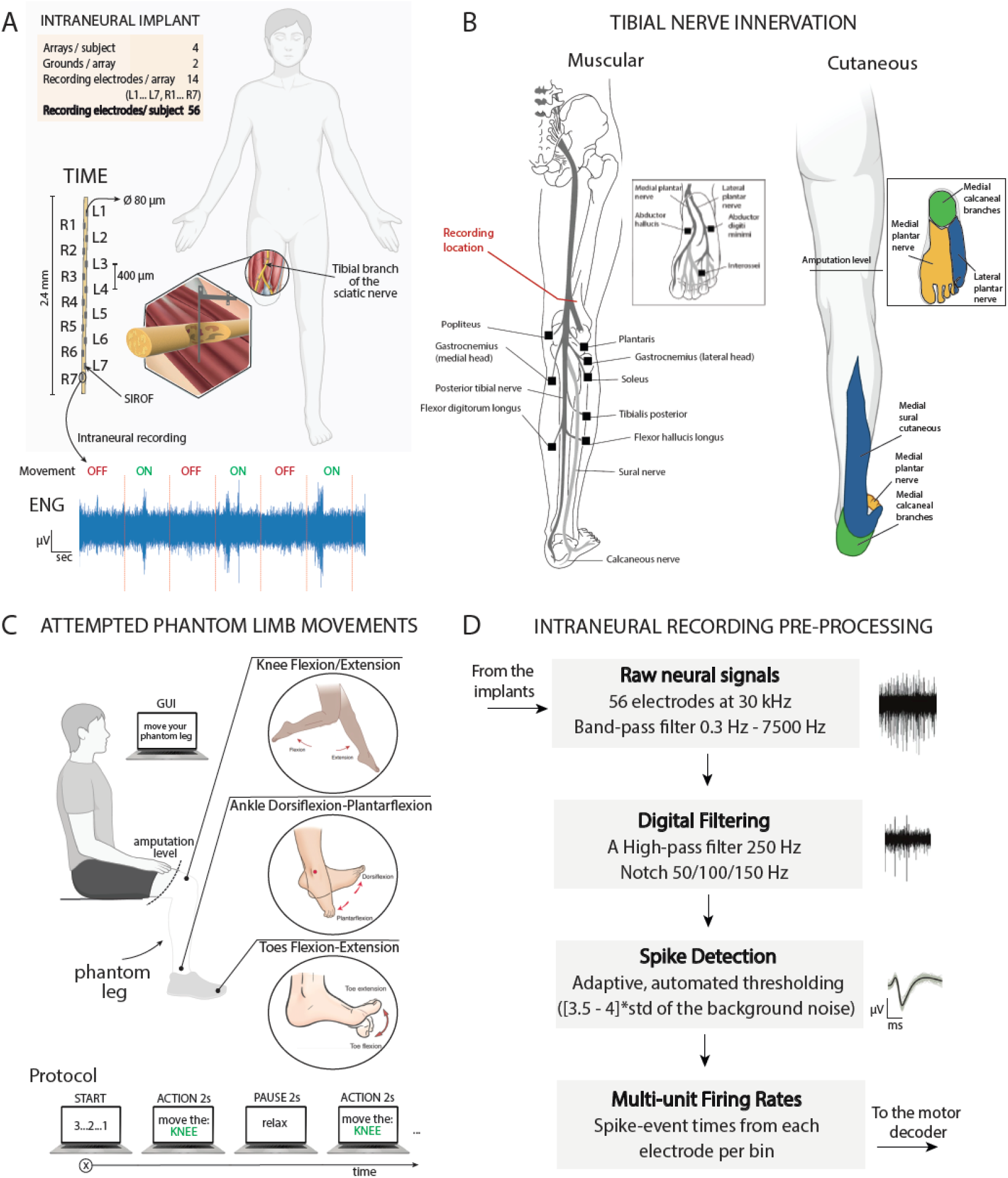
Intraneural recording of the tibial nerve during phantom movements. A) 4 intraneural multi-channel electrodes (TIMEs) were implanted in the tibial branch of the sciatic nerve of two individuals with transfemoral amputation. Each TIME had 14 active sites for recording and stimulation. B) Motor and sensory innervations of the tibial nerve. Implants’ location shown in red. C) Subjects were asked to move their phantom limb following the instructions displayed on a screen. The protocol consists in randomized knee-ankle-foot movements of 2s interleaved with 2s of pause. D) Pre-processing of the recorded ENG signal consisted in a digital filtering, spike detection and multi-unit firing calculation.

The use of SNNs is particularly advantageous for decoding ENG signals, as these networks are designed to process the spiking nature of the data, making them a natural fit for this task. Raw ENG spikes, however, are not sufficient for robust decoding. To improve the signal quality for motor control, we introduce an event-based encoding technique, which converts continuous ENG signals into dense spike trains. This encoding method enhances the signal’s usability by increasing the density of information available for decoding, providing a more accurate and robust system for prosthetic control.

By using dataset recorded from TIMEs and integrating it with SNN-based decoding, and sensory mapping, our work demonstrates a complete bidirectional intraneural interface for lower-limb neuroprosthetics. The use of implantable neurotechnology and neuromorphic hardware^32^ not only for neurostimulation, but also for direct peripheral nerve decoding, paves the way for bidirectional, neurally-controlled prosthetic limbs.

## Results

We recorded residual neural activities from a total of 56 active channels, implanted inside the tibial nerve stump in two AKA patients (**Table S1**), who had been asked to perform three different types of phantom motor tasks (ankle movement, knee movement and toes movements) in two different directions (flexion and extension) each (**Figure 1A-C**). The tibial nerve is responsible for the innervation of the majority of flexor muscles in the lower part of the leg, in particular muscles like the Gastrocnemius, which is accountable for movements of the ankle and knee, as well as the Flexor Digitorum and Flexor Hallucis, instead involved in the flexion of ankle and toes (**Figure 1B**). For this reason, we expected to being able to extract neural modulation concurrently with volitional movements of all the three anatomical regions analyzed, as well as being able to decode the direction of the intended phantom motion.

After pre-process the neural signal, we characterized in detail the neural activity in response to different phantom movements involving multiple joints and directions in both subjects. This neural activation has been then compared to the muscular innervation of the tibial nerve and the specific movement. Then, we design and implemented neural decoders in order to predict the subjects’ intentions to move their phantom limb. We developed SNN-based decoders using the recorded spiking signals as direct inputs, and we compared their performance with that of conventional non-spiking-based decoders. Finally, we expanded the bandwidth of the neural signal to include also the recorded activity of the thigh muscles using the neural interfaces as inter-muscular electrodes. We evaluated the benefits of using hybrid decoders compared to ENG-only decoders.

### The intraneural signal encodes joint-related and direction-related movement of the lower-limb

From the recorded intraneural activity, we observed significant neural modulation associated with movements of the lower-limb (**Figure 2A**). In fact, 91% (51/56 implanted electrodes) of the recording sites showed responsiveness for at least one of the phantom movements analyzed in S1 (**Figure 2C**), with only 5 overall silent electrodes, all located on the same TIME. Of these 51 channels, 90% (46/51 electrodes) showed extension-related neural activation, while 71% (36/51 electrodes) resulted associated with flexion-specific modulation (**Figure 2C**). For S2, instead, the situation seems to be reverted, with only 37.5% (21/56 electrodes) revealing significant neural modulation during the execution of the ankle and toes phantom motions (no data for knee movements is available for S2) (**Supplementary Figure 2A**). Of the active 21/56 electrodes, the 85.7% (18/21 electrodes) shows activation during the flexion phase of the movement, while only 23.8% (5/21 electrodes) of the recording sites seems to be associated with the extension.

**Figure 2.**
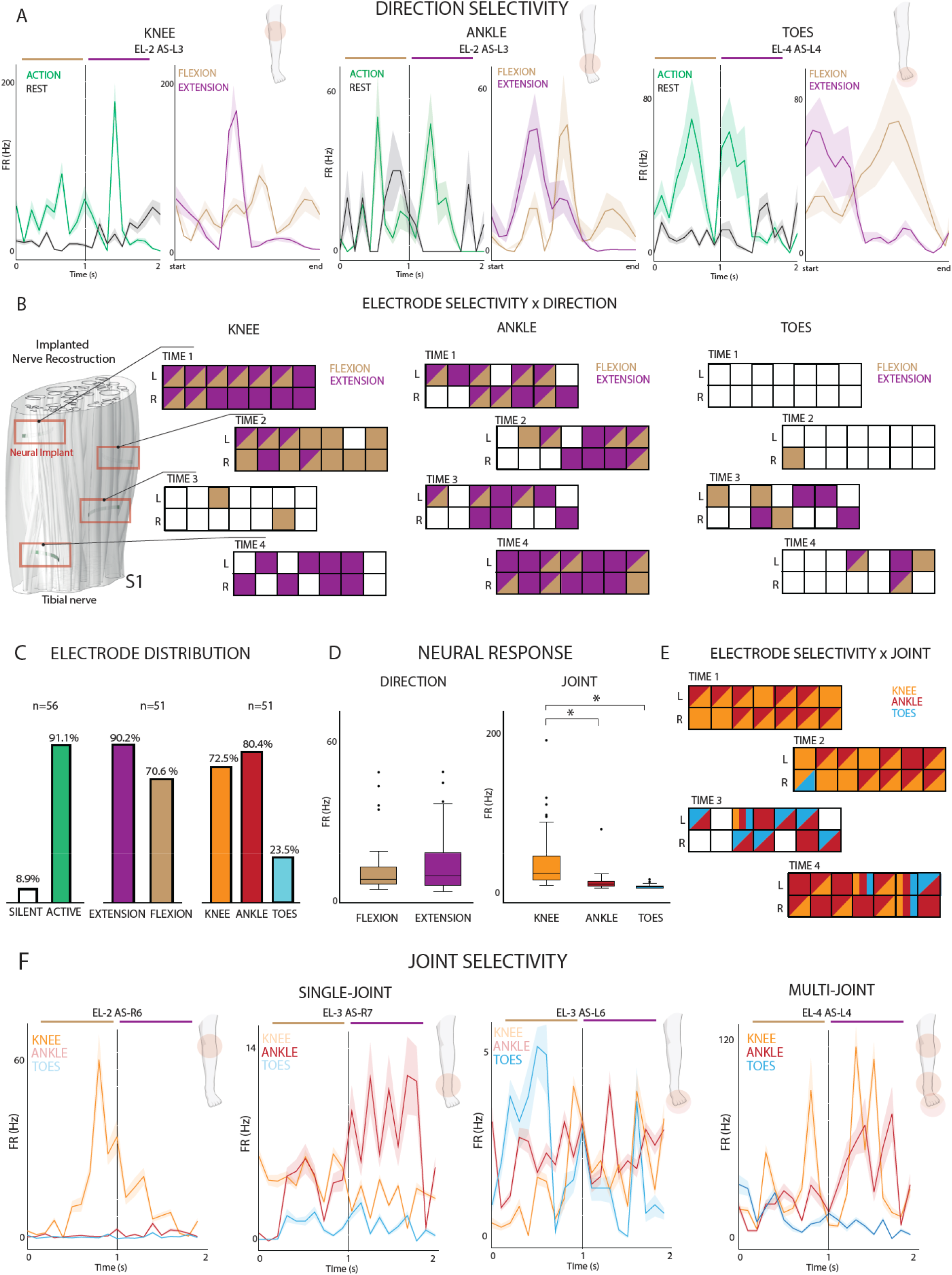
Intraneural recording of the tibial nerve during phantom movements. A) Multi-unit activity of three active sites showing significant direction modulation during flexion (brown) or extension (purple) of the knee, ankle or toes. Mean and SEM are reported. B) 3-D nerve reconstruction of participant S1 with the 4 TIME implanted (left). Each of the 4 TIME are reported for the 3 joint movements, showing the significant modulation for each individual channel. A channel colored in purple shows only significant modulation for flexion, in brown only for extension, both colors both directions and white no modulation. C) Percentage of significant modulation among all 56 channels. Data for silent vs active, flexion vs extension and knee vs ankle vs toes are reported. n indicates the sample size. D) Average firing rates across all channels for each direction and joint involved. On each box, the central mark indicates the median, and the bars the interquartile ranges, IQR. Points indicate the outliers. E) Joint selectivity is reported showing in orange only significant modulation for the knee movements, in red only for ankle and light blue for toes. Mixed colors indicate mixed joint selectivity. A white channel indicates no modulation. F) Multi-unit activity of three active sites showing significant single-joint modulation during knee (orange), ankle (red) or toes (light blue) movements. A channel showing multi-joint selectivity is also displayed (right). Mean and SEM are reported. ^*^p<0.05. Data reported are from S1.

In addition, the firing rates distribution varies across the two directions of the phantom motion. The mean firing rate appears comparable across flexion, 9.58 Hz [interquartile range (IQR), 6.3] and extension, 10.88 Hz [IQR, 12.06] for S1 **(Figure 2D)**. In S2, the mean firing rates computed across only active channels appear much higher and variable for flexion (median equal to 76.35 Hz and IQR = 111.18), while comparable for extension (median = 9.02 Hz and IQR= 22.03).

In S1, the phantom motion of knee and ankle evoked significant neuromodulation respectively in 72.5% (37/51 electrodes) and 80.4% (41/51 electrodes) of the active recording sites, independently from the direction of the motion (**Figure 2C**). Only 23.5 % (12/51 electrodes) revealed neural activation concurrently with the phantom movements of the toes, none of which resulted selectively related to the foot, but always coupled with the ankle joint (7 channels), the knee (1 channel) or both (4 channels). We expected knee motion to drive a higher amount of modulation, being more proximal than the other joints, but we have to consider how the majority of muscles responsible for knee motion are located above the thigh level and are innervated by the sciatic nerve before the branching of the tibial nerve. In S2, the implanted interfaces appear more selective towards nerve fibers related to the ankle phantom motion, with the number of channels reporting significant modulation for the ankle (85.7%, 18/21 electrodes) almost doubling the ones activating concurrently to toes volitional movements (42.8%, 9/21 electrodes) (**Supplementary Figure 2A**). However, from the reported results we can state that the activity was distributed among all the electrodes, showing both single-joint and multi-joint selectivity.

Furthermore, the firing rates associated with the volitional phantom movements of the different joints reveal distinct activation patterns, with peaks at different frequencies (**Figure 2F**). In S1, we observed responses of 10.75 ± 1.02 Hz, 6.80 ± 1.62 Hz and 2.75 ± 0.26 Hz, respectively for knee, ankle and toes movements. The median firing rates were 24.81 Hz [IQR, 30.62] for the knee, 11.07 Hz [IQR, 6.47] for the ankle and 7.19 Hz [IQR, 2.34] for the toes movements, in S1 (**Figure 2D**). The firing rates during ankle and knee movements, as well as knee and toes, are statistically different, while the neural response during ankle and toes is not (Kruskal-Wallis statistical test, n=37 for knee, n=41 for ankle, n=12 for toes, ankle-knee: p < 0.0001, knee-toes: p = 0.0009 and ankle-toes: p = 0.84). Therefore, the peak firing rate ranges seem to be proportional to the anatomical proximity of the phantom joint, consistently with the number of joint-selective channels for S1, even if not completely joint-specific. This is evident also observing the average computed on some example channels in **Figure 2A-F**.

Similar results were observed in S2 (**Supplementary Figure 2B**), where the average peaks patterns computed on active channels only for each joint (in S2 only ankle and toes) are significantly different (Kruskal-Wallis statistical test on the average peaks on active channels, n=18 for ankle, n=9 for toes, p = 0.79). For S2, the distribution of neural response across joints is characterized by median firing rates equal to 59.9 Hz [IQR, 86.75] and 151.0 Hz [IQR, 68.64], with higher peaks associated with toes and overall higher variability with respect to S1.

No significant difference emerges in modulation patterns associated with flexion and extension, concerning the same anatomical region (Kruskal-Wallis statistical test, n=36 for flexion, n=46 for extension, p = 0.78) for S1 (**Figure 2D-A**). Instead, for S2 we observed statistical difference from the firing rates peaks during flexion and extension (Kruskal-Wallis statistical test, n=19 for flexion, n=5 for extension, p = 0.008 for S2).

In addition, for what concerns the spatial distribution across all electrodes (**Figure 2B**), in S1 we noticed a separation between sites associated with flexion phase and extension phase of the phantom motion, in particular in knee and toes, where respectively the 32% and 17% of the modulating sites reveal significant response during both motion directions, while in ankle the overlapping in neural response for the two different phases of the phantom motion interests a wider area of the electrode arrays (46% of the modulating sites, with only 2 electrodes selectively associated with extension). For knee movements, the number of channels modulating during flexion overall is lower than the number of channels modulating for extension, with also a higher number of electrodes selectively associated with extension (15 channels) with respect to the ones selective to flexion (10 channels). Similarly, for the ankle a stronger activation of the recording sites during extension and only 2 channels selective to flexion and 20 selective for extension, while for toes volitional phantom movements, the neural response of electrodes is balanced between the two directions. In S2, the overlapping between modulating sites appears null for the toes, and minimal for the ankle (**Supplementary Figure 2C**). In this last case, only 2 electrodes show significant neural modulation for both directions of the volitional phantom movement, with an evident segregation between channels active for distinct anatomical parts.

### The neural modulation of a specific movement overlaps with its muscular representation

Analyzing the different types of modulation recorded by the 4 electrodes arrays in S1, we can notice how for knee and ankle most channels show significant activation during at least one direction of the motion, with only 33.9% and 26.8% of the total 56 channels being silent during both movements (**Figure 3A**). The opposite, instead, can be observed concurrently with toes movements, where the 78.6% of the recording sites tends to not show any significant modulation during both flexion and extension. While, for toes movements, the number of sites selectively activated only during flexion appears balanced with the number of sites selectively activated only during extension (both 8.9% of the 56 electrodes), the number of sites with significant modulation in both directions is minimal (3.6%). On the other hand, the number of channels selectively activated during flexion were 17.9% and 3.6% (of the total 56 channels) for knee and ankle movements respectively, while those only selective to extension 26.8% and 35.7%. We observed a higher number of channels active during both directions in the phantom movement of the ankle (33.9% of the total 56 channels) compared to the knee (21.4% of the total 56 channels). In S2 (**Supplementary Figure 2**), both for ankle and toes the number of electrodes not displaying any neuromodulation is significantly higher than the amount of active channels (respectively 67.9% and 83.9% of the total 56 channels are silent, for ankle and toes), which also show a prevalence of recording sites selectively associated with flexion direction of the phantom motion (respectively 25% and 14.3% of the total 56 channels, for flexion in ankle and toes, while only 3.6% and 1.8% for extension in ankle and toes). Also, in contrast with S1, in S2 the overlapping in the electrodes evoking neural activity for the two directions of phantom motion is reduced to only two channels for the ankle and null for toes.

**Figure 3.**
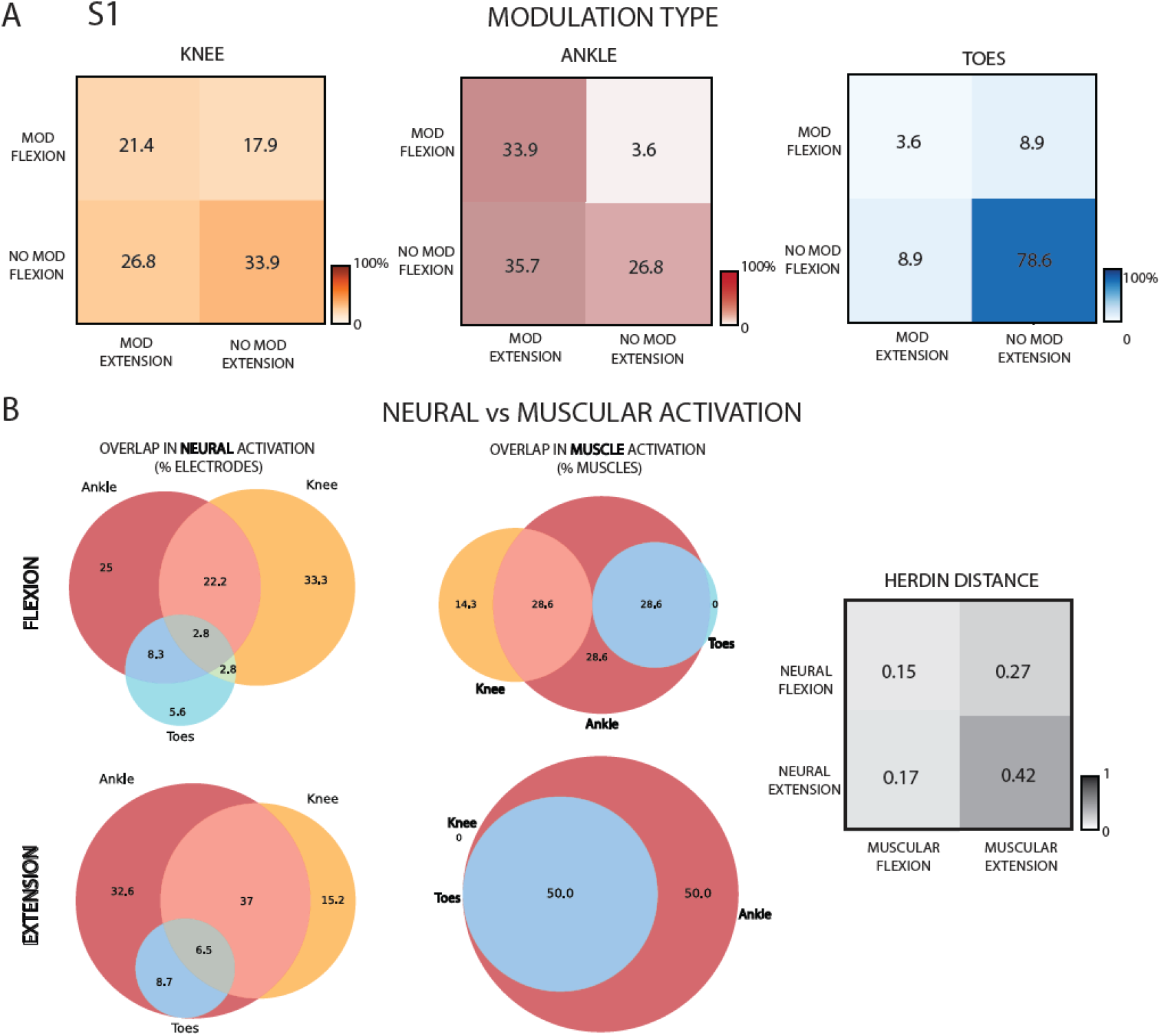
Neural and muscular activations during phantom movements. A) Modulation matrices showing the percentage of electrodes significantly modulating only during flexion, only during extension or both for the three joints. N=56. B) Percentage of electrodes showing a significant modulation in the neural activation for each direction and joint movement (left). Percentage of muscles, innervated by the tibial nerve, involved in each movement. N=56. Herdin distance between the neural activations sand muscular representations for each movement (right). 0=highest, 1=lowest similarity. Data reported are from S1.

To better interpret these modulations, we considered the overlapping of the muscles involved in the different volitional movements of the lower limb. We only included muscles innervated by fibers belonging to the tibial nerve^33^, where the implants were located, as listed in **Table S2**. After extracting Venn’s diagram of the muscular activation and its overlap during phantom movements of the ankle, knee and toes, both for flexion and extension, we transformed them into numerical matrices. We then computed the Herdin Distance between these matrices, representing muscular involvement, and the neural activation, as a metric of similarity [0=highest similarity, 1=lowest similarity]. The results indicated a high correlation between overlapping in muscles participation and neural response during flexion, with Herdin distance equal to 0.15, but a lower similarity between muscular and neural activation during flexion, with an Herdin distance of 0.42 (**Figure 3B**). Instead, surprisingly the correlation between the neural response during extension phase of phantom motion and overlapping in the flexors’ muscles (located in the posterior part of the leg) shows high similarity (0.17). This paves the way to the hypothesis that the recorded neural modulation observed during extension could be partially associated with inhibitory response flowing through the tibial nerve and targeting the flexors muscles^34^, rather than activation impulses directed to their antagonist extensors muscles (located in the anterior lower part of the leg). However, also spatial overlapping in the electrodes neural responses during flexions shows a medium similarity to the overlapping in the extensors’ muscles participation to ankle extension, knee extension and toes extension movements, with an Herdin distance of 0.27. This last result still opens the possibility to have recorded the activity of motor neurons directly innervating extensors muscles.

By cross-referencing the overlapping muscle activations innervated by the tibial nerve (as detailed in **Table S2** and **Figure 3B**), we identified two distinct muscle clusters corresponding to different motor synergies: one involving the ankle-toe muscles (flexor and extensor hallucis longus, flexor and extensor digitorum longus) and another involving the knee-ankle muscles (gastrocnemius and plantaris). These clusters align with the electrode joint-selectivity patterns shown in **Figure 2E**. Specifically, TIME-1, TIME-2, and the left half of TIME-4 are associated with knee and ankle movements, while TIME-3 and the right half of TIME-4 are linked to ankle-toe control. This spatial segregation of electrode selectivity may reflect a physical separation of motor neuron fibers targeting different muscle groups at more distal levels of the nerve. Additionally, muscles such as the soleus and tibialis posterior (ankle) and the popliteus (knee), which also contribute to joint-specific movements and are innervated by the tibial nerve, show selective modulation during phantom movements, further supporting joint-specific functional organization.

### Decoding phantom movements from intraneural recordings using SNN-based decoders

After characterizing the neural efferent response during volitional phantom leg movement conditions, we designed and implemented a SNN to decode motor intent from intraneural ENG recordings. The decoder was trained to classify six classes (ankle flexion/extension, knee flexion/extension, toes flexion/extension) for S1, and four classes (ankle and toes flexion/extension) for S2. The designed decoding architecture used a shallow fully connected SNN composed of fully connected leaky integrate-and-fire neurons (LIF) with synaptic conductance. The input layer comprised 64 neurons, and the output layer had several neurons equal to the number of movement classes. Synaptic transmission was modeled via exponentially decaying conductance. We evaluated this architecture using three different signal encoding strategies. The choice of a single-layer architecture was motivated by the observation that the decoding problem was approximately linearly separable, and deeper architectures did not significantly improve performance, therefore, we favored a compact, low-power design aligned with neuromorphic hardware constraints (**Figure 4A**). All the reported results are indicated as mean test accuracy ± standard deviation (SD) on 5-fold cross validation. All the experiments were run training the network using surrogate gradient descend, with the learning rule indicated as in **Figure 4A**, on 5-Fold cross validation.

**Figure 4.**
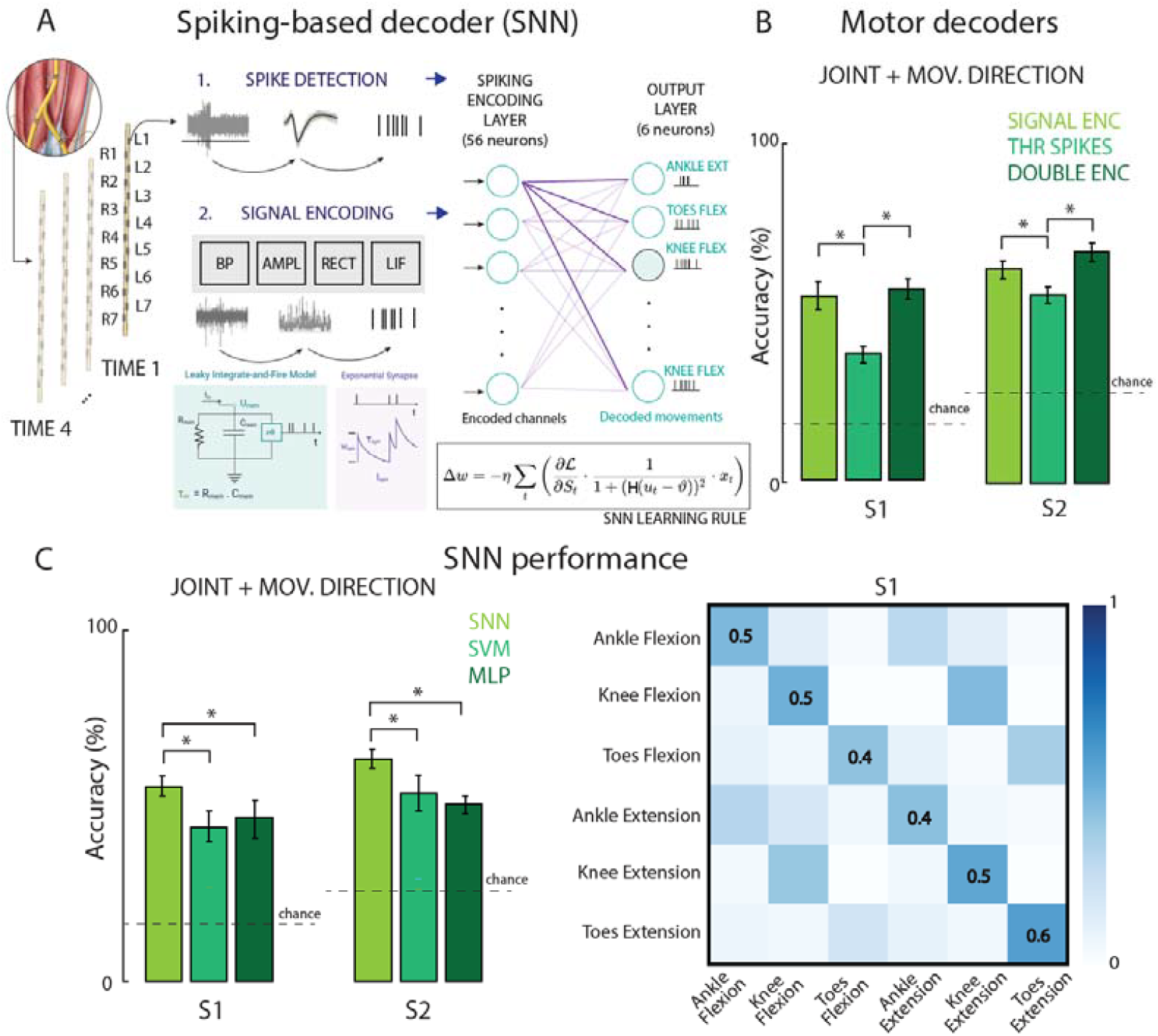
Spiking Neural Networks to decode phantom movements from the intraneural recordings. A) L is the loss (e.g., cross-entropy between membrane potentials and target class), η is the learning rate, u_t_ is the membrane potential at time t, L is the spike threshold, H is the Heaviside function. B) Comparison of the SNN-based decoders’ accuracy in both participants. Performance of the Signal Encoder, Threshold Spikes and Double Encoding are reported. C) Comparison of the decoders’ accuracy in both participants. SNN: Spiking Neural Network, SVM: Support Vector Machine, MLP: Multilayer Perceptron. Error bars indicate mean and STD. Chance levels indicated with dashed lines. ^*^p<0.05. Confusion matrix of the SNN Signal Encoder performance in S1 for all movement types.

First, we evaluated the decoding performance using an encoder that extracted multi-unit activity from the signals through a thresholding method. This first method returned poor results (**Figure 4B**), with 38.86 ±1.67 % of test accuracy on 6 classes for S1 and 55.43 ± 2.77 % test accuracy on 4 classes for S2. Therefore, we tested a more innovative spike encoding method^35,36^, which was based on the elicitation of the membrane sensibility of a very simple LIF neurons with no synaptic conductance, which parameters were fine-tuned offline and outside the model. In this way, we aimed to investigate whether motor information was encoded also in fluctuations of the spike rate or actions potentials, rather than being linked to variation in signal amplitude. The events emitted as output by the encoding LIF layer, after injecting as input current a scaled and rectified version of the filtered ENG signal, extracted features related to the power content of the physiological neural activity and its temporal variation. Using this method, we managed to achieve significant higher performance (paired t-test, p = 0.003 for S1, p = 0.03 for S2), with a test accuracy of 55.14 ± 4.66 % on 6 classes for S1 and 63.46 ± 3.21 % test accuracy on 4 classes for S2. Finally, we also designed a hybrid double encoding, concatenating the two above-mentioned encoding in a double dimensionality representation for each data sample. In order to be classified, this last encoding required an adjustment of the previous architecture, going from 64 to 112 neurons in the input layer. The achieved results were statistically higher than the ones obtained using the threshold encoding method (paired t-test, p = 0.0012 for S1, p = 0.001 for S2), with test accuracies of 58.29 ± 3.31 % for S1, on 6 classes, and 68.29 ± 3.88 % for S2, on 4 classes. Instead, its performance was not significantly higher (paired t-test, p = 0.18 for S1, p = 0.06 for S2) with respect to using the LIF encoding method, that instead required a network with one-half connections weights to be learned, being therefore more memory efficient.

We compared the results achieved by the SNN decoder on the LIF encoded neural data with two types of more conventional machine learning tools, like a linear Support Vector Machine (SVM) and Multi-layer Perceptron (MLP), with a hidden layer of 236 non-spiking artificial neurons (**Figure 4C**). These methods are commonly used as baseline classifiers in many computational neuroscience studies as motor decoders^37^. Both SVM and MLP were firstly tested using variety of signal features, such as spike rate, root mean square (RMS) and power (**Supplementary Figure 3**) to find the best possible. For both classifiers, RMS and power were the features achieving higher decoding performance (not statistically significant with respect to each other. Paired t-test, p = 0.39 for MLP, p = 0.79 for SVM for S1; p = 0.49 for MLP, p = 1.0 for SVM for S2), while spike rate performed quite poorly in both S1 and S2 (Paired t-test, p = 0.0004 for MLP, p = 0.03 for SVM for S1; p = 0.0001 for MLP, p = 0.0003 for SVM). Since the LIF encoding, as explained above, tends to extract features related to frequency power from the signal, we decided to compare the results achieved by the SNN with the performance of SVM and MLP using power as feature (test accuracy equal to 43.70 ± 5.19% on 6 classes for S1 and 53.75 ± 5.43 % test accuracy on 4 classes for S2 for SVM; while test accuracy equal to 46.85 ± 6.48 % on 6 classes for S1 and 50.83 ± 3.41 % test accuracy on 4 classes for S2 for MLP. No significant difference between SVM and MLP accuracies: paired t-test, p=0.44 for S1, p=0.55 for S2). The SNN achieved significantly higher performance with respect to both SVM and MLP (**Figure 4C**; paired t-test, p = 0.04 for SNN vs MLP, while p = 0.0029 for SNN vs SVM for S1; p = 0.006 for SNN vs MLP, while p = 0.018 for SNN vs SVM for S2). Notably, a sparser data representation and computation, which also mimics the natural physiology of information transmission inside biological nerves, results in a better processing of data and overall better efficiency in terms both of decoding performance and memory/energy usage.

Finally, by analyzing the confusion matrix returned on the whole test set by the SNN trained on LIF encoded spike arrays (**Figure 4B**) for S1, we can appreciate how the decoder seems able to correctly identify the 6 classes with accuracies that range from 41% to 57% of accuracies. Still the network sometimes mistakes the flexion phase of a specific phantom motor task with the extension direction of the same (38%, 44%, 34% of the time respectively for ankle, knee and toes). Or vice versa, it misleads extension execution of the motion with its flexion counterpart, with the same frequency for ankle (30% of the time) and slightly less frequently for knee and toes (39% and 19% of the time respectively). This could be due to neural activation overlapping or variable timing in the execution of the volitional imaginary movement (**Figure 3B**). Instead, the errors between different joint-related movements are limited (between 0 and 11%), with the only exception of the extension of the ankle, which gets mistaken for the flexion of the knee 17% of the time.

In S1, overall better accuracies reached for extension are in line with the spatial activation and selectivity of the neural response of the electrodes (**Figure 2B**). In fact, having more recording sites selective to extension movements than to flexion-only leads to a 10% difference in test accuracy when distinguishing between the two phases of the same motor task. Surprisingly, despite the low level of recording selectivity in toes phantom movements, toes extension results the class for which the network reaches the highest performance, setting an unprecedented achievement in the neural decoding of the lower-leg movement.

### Informing neural decoders with muscular-related signals improves their performance

While processing the ENG data, we filtered the neural signal in a bandwidth between 250 and 7500 Hz as already reported in previous studies^30^. In addition, since the electrodes were implanted in the nerve and located in between the thigh muscles, we also decided to extract inter-muscular (imEMG) information from the recorded activity by filtering the signal between 50 and 350 Hz, frequencies normally related to EMG activity^38^.

In S1, we observed a significant modulation in the neural activity, in particular associated to the ankle and knee phantom motion (**Figure 5A**). Giving this imEMG signal as input to our SNN decoder, a performance of 63.14 ± 6.15 % on 6 classes for S1 and 72.29 ± 3.68 % test accuracy on 4 classes for S2 were obtained (**Figure 5B**). Compared to the information contained solely within the ENG-related frequency band (350–7500 Hz) — which yielded a test accuracy of 51.14 ± 5.06% across 6 classes for S1 and 64 ± 1.07% across 4 classes for S2, after filtering out all other frequencies potentially associated with residual muscular activity — the decoder trained on imEMG frequency range achieved significantly higher performance (t-test, p = 0.024 for S1, p = 0.023 for S2). For this reason, we decided to inform our decoders with also muscular signals, by considering the whole frequency range going from 50 to 7500 Hz (ENG+imEMG). The ENG+imEMG resulted in a higher decoding performance than when using ENG frequency band only (**Figure 5B**). The increases were +13.2% for S1 and +8.6% for S2 (test accuracy of 64.29 ± 3.26 % on 6 classes for S1 and 72.57 ± 2.10 % test accuracy on 4 classes for S2; t-test, the increment is not statistically significant for S1: p = 0.06, while for S2: p = 0.003). The performance was not significantly higher when only imEMG band was chosen (t-test, p = 0.77 for S1, p = 0.89 for S2). With respect to the results achieved using only the ENG bandwidth (350-7500 Hz), including the lower frequencies associated with residual muscular activity results in an improved ability to correctly identify the phantom movements of ankle and knee (**Figure 5C**). Specifically, the network leverages the imEMG signals from muscles involved in ankle and knee movements to better capture the motion patterns of these anatomical regions and more accurately decode the intended direction of movement. The drawback is a slight decrease in the ability to correctly identify the toes movements, now characterized by much lower amplitudes with respect to the other classes.

**Fig. 5.**
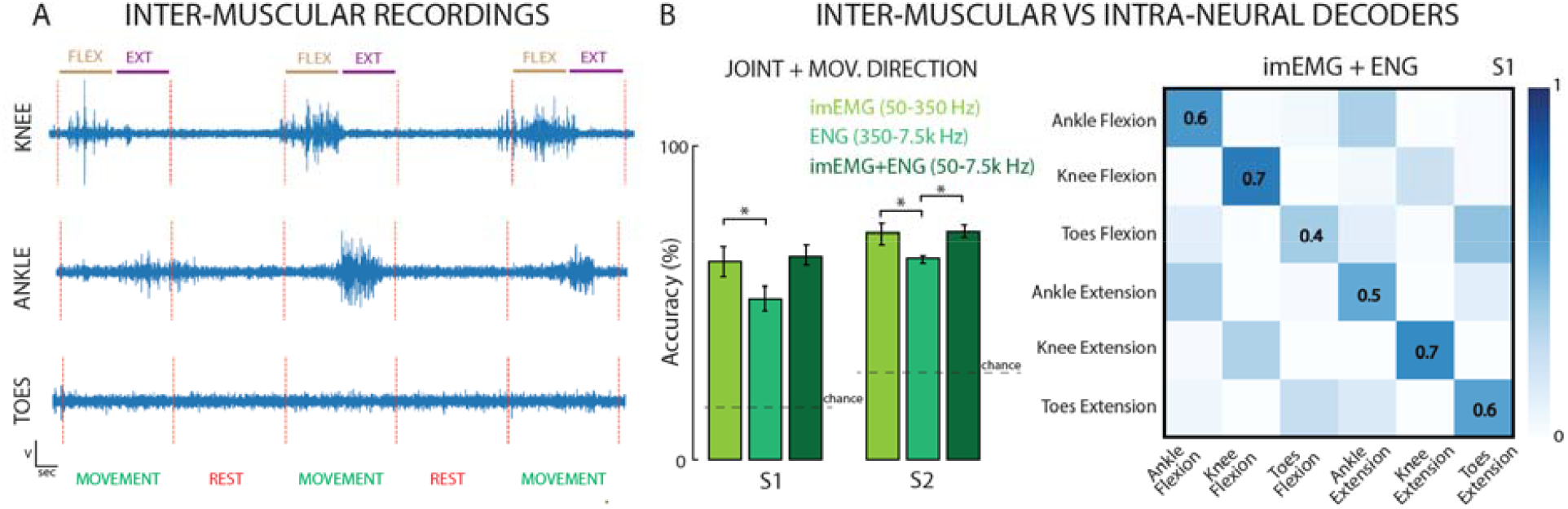
Informing intraneural decoders with inter-muscular EMG signals. A) Example of inter-muscular signals filtered between 50 – 350 Hz. B) Comparison of the SNN-based decoders’ accuracy in both participants using imEMG, ENG or imEMG+ENG bandwidths. Error bars indicate mean and STD. Chance levels indicated with dashed lines. ^*^ t-test, p<0.05. Confusion matrix of the SNN Signal Encoder performance in S1 using the imEMG+ENG bandwidth (50 Hz - 7.5 kHz).

We proved the possibility to use nerve interfaces to record not only neural activity related to phantom movements, but also imEMG activation of the residual and above-amputation level muscles, which can be exploited to improve the performance of the motor decoder also in the absence of the limb.

### Intraneural electrodes allows to record efferent activity and to stimulate afferent fibers

In previous studies on the same patients and using the same intraneural implants^22,23^, we showed that it was possible to elicit physiologically-plausible sensory feedback after delivering electrical current with variable intensity, duration and frequency via the implanted TIMEs.

In S1, by delivering current from all the 56 TIME sites independently, it was always possible to evoke artificial percepts (**Figure 6A-B**). However, only 12.5% of the electrodes (7 electrodes) targeted sensations related to the calf and knee, and 26.7% (15 channels) perceptions associated with the posterior ankle and heel. Interestingly, 60.7% (34 electrodes) of the total recording sites, instead, evokes tactile sensations associated with the toes and upper part of the sole. For all three anatomical parts, the overlap between evoked-sensation and significant neural modulation sites is minimal (7.1% for knee, 16.1% for ankle and 7.1% for toes/foot of the 56 total recording sites) and, overall, lower than the number of channels not targeting any efferent nor afferent fibers (28.7% for knee, 16.3% for ankle and 25% for toes/foot of the total 56 channels).

**Figure 6.**
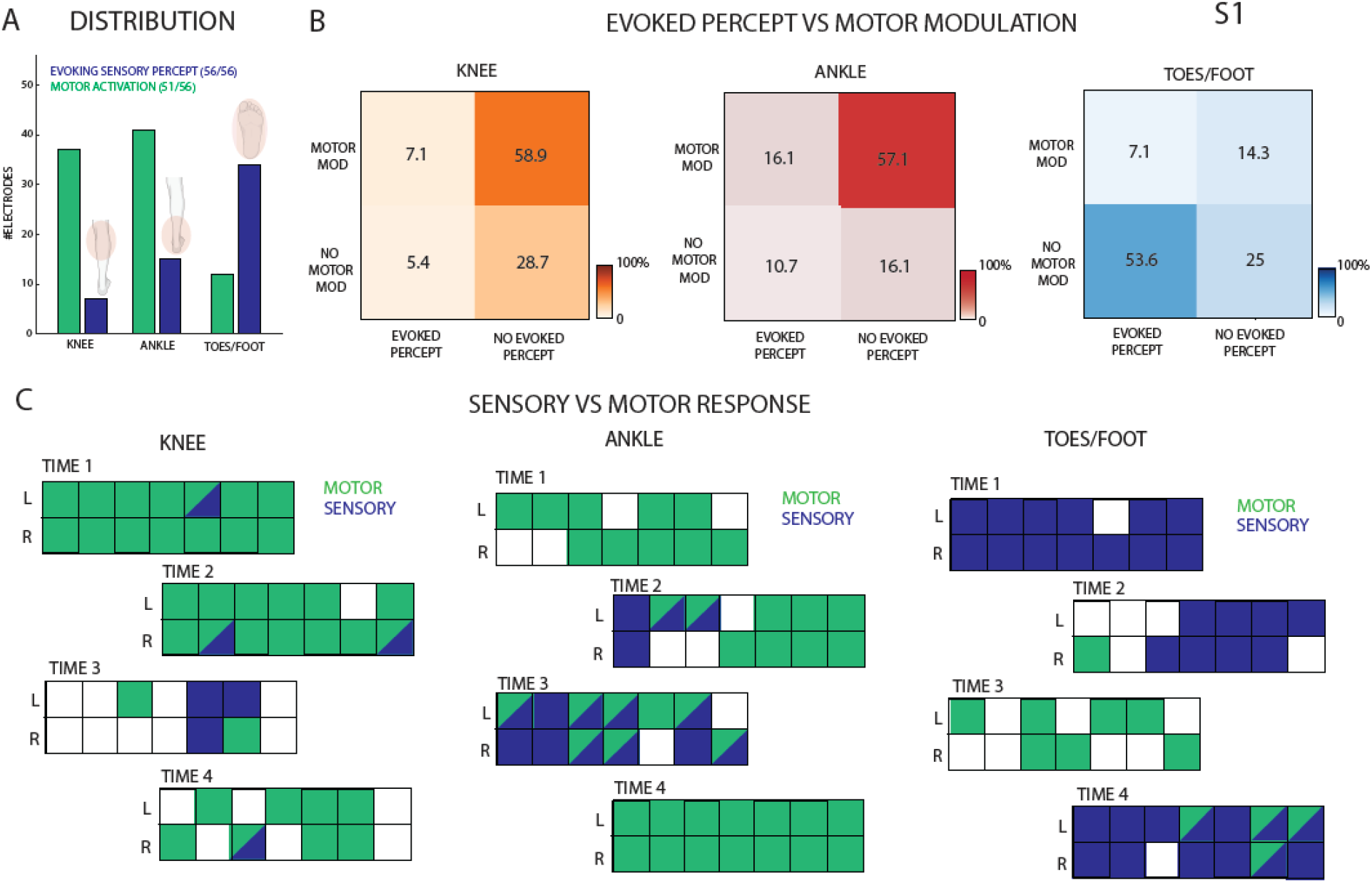
Motor modulation and sensory restoration. A) Distribution of the electrodes showing significant motor modulation compared to those evoking sensation in the knee, ankle or foot areas. B) Modulation matrices showing the percentage of electrodes significantly modulating only during movement, evoking only sensations or both for the three joints. n=56. C) Each of the 4 TIME are reported for the 3 joints, showing the significant motor modulation (blue) or sensory evoked response (green) for each individual channel. A channel colored in blue shows only significant motor modulation, in green only for evoked-sensations, both colors both responses and white no response. Data from S1.

However, while a large proportion of electrodes were responsive solely to volitional phantom movement of the knee/calf (58.9%) and ankle/heel (57.1%) regions—compared to a much smaller fraction located on afferent fibers targeting the same areas (5.4% for the knee and 10.7% for the ankle)—the opposite trend was observed for the toes/foot, where afferent response was more prevalent (**Figure 6A**). In fact, 53.6% of the total 56 sites evokes sensations on the sole and toes, while only 14.3% appears able to record activation during attempted flexion and extension. The low motor selectivity of the recording sites is also associated with a clear spatial separation between electrodes recording efferent motor neurons with respect to channels implanted close to afferents somatosensory fibers (**Figure 6C**). This is particularly evident for the foot, but also true for ankle/heel and knee/calf where the overlapping between motor selective electrodes and active sites eliciting sensations is limited. This suggests that the afferent and efferent fibers in the tibial nerve result to be already segregated within the nerve at the implantation level, before the bifurcation of the sciatic nerve into tibial and common peroneal (**Figure 1B**).

The spatial separation between sites selective to afferent and efferent fibers is confirmed also in S2 (**Supplementary Figure 4B-C**), with an overlapping of only 12.5% and 5.3% of the total 56 recording sites, respectively for ankle and toes. The distribution of the electrodes eliciting percepts appears still mirroring the one seen in S1, with the number of sites associated to sensations of the foot/sole higher than the ones related to the ankle/heel (**Supplementary Figure 4A**). In this case, not all the electrodes if independently stimulated were able to evoke a percept (35/56). Additionally, 60.7% of the 56 channels for the ankle/heel region and 48.2% for the toes/foot region are not involved in either evoking sensations or recording motor modulation (**Supplementary Figure 4C**).

## Discussion

### Intraneural interfacing allows the access to intrinsic muscles signals

In this study, we demonstrated the feasibility to record movement-related information from signals recorded from inside the leg nerves. The information both direction-related and joint-related was captured by the implanted electrodes. Given the ability of the amputees to generate motor activity associated with attempts to control a missing limb movement within a short period of time, we infer that the motor central and peripheral pathways remain largely intact from the functional point of view, as shown by imaging studies^39^. This is in line with what observed on the sensory pathways, given the easily identified, discrete and graded, sensations elicited by direct electrical stimulation of the nerve^22^, without the need for any sensory training.

The use of 4 TIMEs in a single nerve with a diameter of around 1 cm allowed the recording of significant modulation for each of the performed movements in both participants (S1: knee-ankle-toes, S2: ankle-toes). This result indicates an appropriate coverage of the cross-sectional nerve space with this number of penetrating multi-channel electrodes. Indeed, not all the joints’ movements were recorded on every single electrode (e.g., in S1 TIME-1 and TIME-2 showed modulation during knee and ankle movements, while toes-related movements were detected mostly on TIME-3 and TIME-4) supporting the appropriate selectivity with 4 implants. This electrode placement has also been suggested for stimulation purposed using realistic computer simulations^40,41^, considering the nerve fascicular topography^42^.

The type of modulation response, observed in the implanted nerves, was mostly an increase of the multi-unit nerve activity (firing rates) locked to the phantom movement onset or offset. The average aggregate firing rate was comparable to that reported in studies of upper-limb nerve intraneural recording^26,30^ in humans during hand movements. However, in case of more distal joint-related movements we observed a decrease in nerve firing possibly due to the proximal electrode location (thigh level) and to the different number and size of the muscles, generated force, and number of recorded efferents, involved in that isotonic movement^43,44^.

In addition, thanks to the intimate contact of this penetrating neurotechnology with the nervous fibers, it was possible to capture the neural modulation in response to different attempted movements of the phantom limb in both amputated individuals. Specifically, a significant modulation was observed, not only for proximal limb movements, such as knee flexion and extension, but also to foot-related movements. Interestingly, movements of the toes are controlled by intrinsic leg and foot muscles, such as the flexor hallucis longus (for flexion of the great toe) and flexor digitorum longus (for flexion of toes 2-5), that are lost after a so proximal limb amputation. This is also the case for muscles controlling the ankle flexion (also known as dorsiflexion) and extension (plantarflexion), such as the plantaris and soleus (**Table S2**). This reported evidence shows the potential of such an approach that allows to access to information, not available to non-invasive^45^ or other surgical approaches targeting only residual muscle signals^46^.

### Spiking-based decoders outperforms conventional decoders for neural signals

In this study, we implemented and tested SNN-based decoders able to extract motor intent from intraneural ENG recordings. The SNN significantly outperformed conventional non-spiking decoders. The performance advantages stem from their ability to leverage the temporal precision and discrete nature of action potentials, which carry rich, high-fidelity information about neural intent. Unlike conventional decoders that typically rely on averaged firing rates or continuous signal features (e.g., RMS or low-frequency envelopes), spiking-based approaches preserve the fine-grained timing of neural events.

This allows them to capture rapid changes in neural activity and subtle variations in motor commands that are essential for precise and responsive control. Furthermore, spiking activity tends to be more robust to noise, and drift compared to low-frequency field potentials^47^. These properties make spiking-based decoding particularly well-suited for applications in neuroprosthetic control, where both accuracy and responsiveness are critical. Moreover, our encoding method, also based on LIF neuron dynamics, enhanced the separability of ENG signals by transforming raw neural activity into event-based spike trains. This encoding leverages the integrative properties of spiking neurons to emphasize temporally stable features while attenuating high-frequency noise—critical for handling the low-SNR conditions typical of intraneural recordings. The synergy between spike-based encoding and spike-based decoding results in a coherent, noise-resilient pipeline that improves classification robustness.

The decoder was intentionally designed as a shallow, single-layer network of LIF neurons. This choice was driven by two key observations: (i) the decoding task was approximately linearly separable, as confirmed by the strong performance of linear SVMs, and (ii) deeper networks, while theoretically more expressive, tend to overfit and generalize poorly in small datasets^48^. The single-layer architecture offered an optimal trade-off between complexity and generalization, achieving high decoding accuracy with minimal training data and parameter overhead. Additionally, the proposed SNN-based decoder is also well-suited for implementation on neuromorphic hardware, which mimics neural architectures and processes spikes natively^32^. This makes it feasible to embed the entire decoding process directly into a prosthetic device, enabling real-time, onboard motor control without the need for external computing systems. Such integration not only improves the usability and autonomy of neuroprosthetic systems but also brings them closer to clinical viability. Overall, the use of a compact, biologically inspired SNN decoder enabled accurate and efficient decoding of volitional lower-limb motor intent from intraneural signals, highlighting the potential of neuromorphic approaches in developing next-generation bidirectional neuroprostheses.

### Implantable intraneural electrodes allow for a bidirectional neural interfacing

The here reported intraneural electrodes showed to enable a bidirectional interfacing with peripheral nerves. We demonstrated to successfully decode attempted movements from the recorded signals, and to evoke informative tactile sensations in lower-limb amputees^23^. The recorded neural modulation was mostly in response to knee and ankle movements, while less channels showed modulation during phantom foot movements. On the other hand, the sensations evoked by directing stimulating the nerves were more often reported on the plantar area of the foot compared to the lower leg. This result respects the muscular and cutaneous innervations of the tibial nerve (**Figure 1B**). Indeed, more afferent fibers innervate the plantar region of the foot^49^, and thus it will be easier to activate them in the nerve, evoking a sensation on the foot. Considering their placement within the tibial nerve, we observed limited cluster and somatotopic organization of motor and sensory fibers at the thigh level. This is in line with analyses on the fascicles distribution within somatic nerves, in which the proximal segment corresponds to a region where, as demonstrated by ChAT+ immunostaining, motor axons are heterogeneously distributed and no prediction of their destination can be established a priori^50^.

Interestingly, thanks to the location of the implants at the thigh level, this neural interfacing allowed the recording of both neural signals, and also EMG-related signal, working as inter-muscular electrodes. Similar EMG spiking has previously been reported for intraneural recordings in individuals with upper-limb amputation^51^. To maximize the decoding performance, we showed that including the band of the imEMG signal could potentially improve the decoding accuracy allowing for a more stable control of multiple DOFs, avoiding additional surgical intervention as TMR, AMI or RPNIs^8^.

In the context of bionic legs, this direct interfacing using TIMEs holds significant promise for allowing the bidirectional neural connection between the human peripheral nervous system and a prosthetic device. This would potentially support: i) the restoration of both high-resolution tactile feedback and direct neural control; ii) a direct access to intrinsic muscular information and thus to extract all the volitional movements of the users; iii) a single implanted technology. This might improve not only prosthesis functionality and control accuracy, but also device embodiment and the overall user experience in individuals with limb loss.

### Limitation of the study

There are some limitations to this study that should be considered. First, the small sample size limits how broadly the findings can be applied. While the results are encouraging, more participants would be needed to confirm that the approach works consistently across individuals and amputation levels especially given the variability in nerve anatomy, implantation and neural signal quality.

We cannot extrapolate from these findings what would occur over longer periods of time when trying to decode phantom movements. Over time, signal quality can decline due to factors like tissue response around the electrode, slight shifts in position, or nerve adaptation, as already shown for TIME in humans^52,53^. Studies specifically devoted to this issue are required and are well beyond the scope of the experiments reported here.

Additionally, the electrodes were implanted in only a single peripheral nerve (tibial), which constrains the amount of motor information that can be accessed. While this simplifies the procedure and reduces risk, it also limits the potential for more complex or naturalistic movement decoding, and ultimately prosthetic control. Implanting both the tibial and the peroneal breaches of the sciatic nerve could potentially expand the repertoire of decodable limb movements.

Finally, although we demonstrated motor decoding, this was done in an offline environment. Real-time use would introduce more variability and noise, which could challenge the decoder’s performance. Moreover, the use of the same electrodes also for delivering neurostimulation to restore sensory feedback would introduce stimulation-artifacts. Techniques based on blanking^54^ or time-division multiplexing^55^ should be implemented to allow a stable bidirectional configuration. Future work should explore how the neural decoding holds up under more dynamic and bidirectional conditions.

## Methods

### Participant recruitment and surgical procedure

Two unilateral transfemoral amputees were included in the study. All of them were active users of passive prosthetic devices (Ottobock 3R80) (**Table S1)**. Ethical approval was obtained from the institutional ethics committees of the Clinical Center of Serbia, Belgrade, Serbia, where the surgery was performed (ClinicalTrials.gov identifier NCT03350061). All the subjects read and signed the informed consent. During the entire duration of our study, all experiments were conducted in accordance with relevant EU guidelines and regulations.

Four TIMEs^56^ (14 active sites each) were obliquely implanted in the tibial branch of the sciatic nerve of each subject. The surgical approach used to implant TIMEs has been extensively reported elsewhere^57^. Briefly, under general anesthesia, through a skin incision over the sulcus between the biceps femoris and semitendinosus muscles, the tibial nerve was exposed to implant 4 TIMEs. A segment of the microelectrodes cables was drawn through 4 small skin incisions 3 to 5 cm higher than the pelvis ilium. The cable segments were externalized (and secured with sutures) to be available for the transcutaneous connection with a neural stimulator. After 90 days, the microelectrodes were removed under an operating microscope in accordance with the protocol and the obtained permissions.

This study was performed within a larger set of experimental protocols aiming at assessing the impact of the restoration of sensory feedback via neural implants in leg amputees during a 3-month clinical trial^22–25,57^. The data reported in this manuscript was obtained in multiple days during the 3-months trial in two leg amputees.

### Intraneural electrodes and recordings

Each of the TIMEs (latest generation TIME-4H) implanted in the two amputees was constituted by 14 active sites (AS) and two ground-electrodes. Details concerning design and fabrication can be found in ^58^. For each subject, 56 electrode channels were then accessible for recording and stimulation on the tibial nerve^52^. The neural recorder was the Grapevine neural Interface System (Ripple, LLC), which is a commercial device that can be used for the recording of neurophysiological data through up to 64 high-impedance microelectrodes, divided into 2 ports of 32 poles each. Therefore, up to 4 electrodes (56 active sites) can be recorded simultaneously and digitally sampled at 30 kHz, and each port addressed two TIMEs. Each electrode includes 14 capacitively coupled active outputs (channels - AS) and 2 non-capacitively coupled references.

### Experimental protocol and movement tasks

The participants were sited, and they were asked to attempt movements of the knee, ankle and toes with their phantom limbs, as shown on a screen. The type and the timing of the task (start time, and end time) were specified using a real-time custom interface designed in MATLAB (R2016a, The MathWorks, Inc., USA) positioned in front of the participants. We asked the participants to performed six different limb movements: Knee Flexion, Knee Extension, Ankle Dorsiflexion, Ankle Plantarflexion, Toes Flexion and Toes Extension (**Figure 1C**). Each movement was randomized in 3 blocks and repeated 10 times. The subjects had to move their phantom limbs for each trial as required; one trial lasts 2 seconds and was followed by 2 seconds of rest (no movement). The total movement consists of 1 second for the flexion and 1 second for extension. Here we reported recordings in 2 sessions for S1, 13 and 90 days after implantation and 1 session for S2, 90 days after implantation. Data recordings of S2 during the knee movements were found to be corrupted during the registration process and were therefore excluded from the analysis.

### ENG pre-processing analysis

Collected ENG data were first analysed in MATLAB (R2024a, The MathWorks, Inc., USA) and then processed using Python (version 3.11.9, the Python Software Foundation) in the Visual Studio Code (VSCode) environment. We applied the detailed analysis offline, as following:

#### Corrupted file removal

Firstly, we manually discarded some of the recorded files, since they were associated with flat, cut paths or massive artifacts. In particular in S2, data related to knee movements were found to be corrupted during the registration process and were therefore excluded from the analysis.

#### Temporal Filtering

Raw ENG data from 56 active sites were pre-processed with by applying first an infinite impulse response (IIR) notch filter at the multiples of 50 Hz (between 0 and 10 kHz) with a quality factor of Q=100, to remove any interference due to power line-effect, which are prone to appear at periodical frequencies. We then removed signal drift and non-neural frequency content by designing and using a 4^th^ order Butterworth filter band-pass filter between 250 and 7500 Hz, in accordance with preceding studies processing intraneural recordings in upper-limb amputees^30^.

#### Inter-muscular activity

To assess the amount of information related to the imEMG activation in the recorded signal, we substituted the temporal filtering in the previous step with at 4^th^ order Butterworth filter band-pass filter between the EMG-specific bandwidth 50 and 350 Hz. We called the so filtered neural signal imEMG path. We also compared the classification performances achieved on the imEMG signal with a version of the ENG signal filtered between 350 – 7500 Hz, so by only considering the information related to the neural activation and completely discard the muscular residual content (called ENG in **Figure 5**). Finally, we also considered the signal recorded filtered on the total bandwidth (between 50 -7500 Hz), including both inter-muscular residual activity and modulation associated uniquely with the axons, therefore called imEMG+ENG.

### Characterization of intraneural spiking activity

#### Spike detection

To characterize the evoked neural activity, as an electrode-specific metric of the modulation during motion as opposed to the inter-actions resting condition, we performed a multi-unit spike detection, involving a simple subject-adaptive, automated thresholding. After a keen tuning, the threshold value was set equal to a factor, respectively to 3.5 and 4 for S1 and S2, multiplied by the standard deviation of the signal^59^. The threshold was determined separately for each recording channel.

#### Motor maps extraction

From the obtained spike-events times from each electrode, we drew the firing rates by summing the number of spikes appearing in non-overlapping temporal bins of 100 ms duration (**Figure 1D**). As a quantitative metric of the neuromodulation, for each electrode we computed the z-score, as the difference between the mean firing rates during action 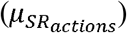 vs rest 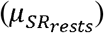 divided by the standard deviation of the firing rate during rest 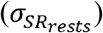:

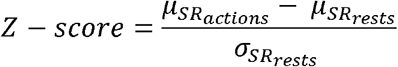

If the z-score exceeded the value of 0.5, we assumed neuromodulation associated to a specific phantom motion present in the considered site.

### Spike-based encoding

We tested a method to encode the information contained in the analog signals into spike arrays, to be input into the SNN encoder, based on the elicitation of the membrane sensibility of a simple LIF neuron. This method was implemented on simulation using the Brian2 software^60^. Before providing the pre-processed signal to the neuron, it was full-wave rectified, in order to preserve both the positive and negative signal phases, and scaled using z-score normalization. The dynamics of the membrane of this neuron can be modelled as:

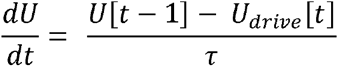

Where *U* is the LIF neuron membrane potential, *U*_*drive*_ is the continuous domain ENG signal, acquired by a specific channel, and *τ* is the time constant of the membrane. If no input is injected into the neuron, the membrane potential tends to decay in an exponential way in time, with time constant *τ =* 10 *ms* , till it reaches its resting state value. If the membrane of the neuron reaches a specific threshold value (in our case set to *thr* 350 *mV*), the neuron emits a spike/event as output and the membrane potential is reset to a specified value (*U*_*reset*_ = 0 *mV*). The simulation was carried out for the entire duration of the recording, and we employed a refractory time of 1 ms, in which the LIF neuron cannot spike even if the threshold is reached. This type of event-based encoding extracts information related to the power contained in the signal, and its rate of variation in time in a biologically-inspired way that resembles how the human body processes sensory information (like for example in the cochlea^61,62^). This encoding was later binned using 2.5 ms non-overlapping windows (the same length of the simulation time step), in order to reduce the dimensionality of the generated event-representation.

### Conventional motor decoders

We employed two traditional machine learning classifiers as baseline motor decoders: a SVM with linear kernel and an MLP. At first, we also tried implementing an SVM decoder with radial basis function (RBF) kernel, to assess it the translation of the data in a non-linear alternative feature representation could improve classification, but we eventually realized that the linear SVM always outperformed the one with RBF kernel, which was then discarded for further analyses. Instead, we designed MLP with 112 neurons in the input layer and 236 (210% of the number of input neurons) in the hidden layer. The output layer dimensionality was set to 6 for S1 and 4 for S2, in compliance with the classification task.

### Feature extraction for conventional decoders

The SVM and MLP classifiers were trained on signal segments on 100 ms as input data samples. For each data sample, we computed the following features, separately for each channel:

#### Root Mean Square (RMS)

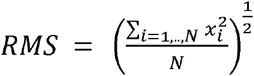

#### Power

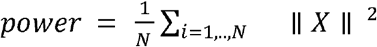

*Spike Rate (SR)*: as the sum of spikes (obtained using the same thresholding method seen for the characterization and quantization of the spike activity) normalized on the window length.

Each of these features were computed on temporal windows of 50 ms. Then, the two values obtained independently for each electrode were concatenated in a single feature-vector representation of the data sample ∈ ℝ^112^.

### Shallow Spiking Neural Network (SNN)

We designed a single-layer SNN comprising 56 input neurons (or 112, when processing the signal represented using the double encoding, which combined both the thresholding and the LIF-neuron encodings) densely connected to the output layer. The output layer consists of 6 for S1 or 4 for S2 LIF-neurons, encoding the classification assignment for each data sample. Neurons receive the encoded spike trains from the 56 ENG channels, once removed the grounds. The network is designed and trained using SNNTorch^63^ (version 0.9.1), a python package that extends the capabilities of PyTorch (version 2.3.1), taking advantage of its GPU accelerated tensor computation and applying it to spiking neurons. Neurons are simulated as second-order LIF neuron model, using the Synaptic class already available on SNNTorch, which considers also the synaptic conductance, modelling the synapses between neurons as exponential kernels.

When an input spike arrived, the synaptic current is incremented by the corresponding synaptic weight. Subsequently, the synaptic current decays exponentially over time with a time constant denoted by *τ* _*syn*_. The neuron integrates the incoming synaptic currents in the membrane potential *U* , which instead leaks over time with a time constant *τ*_*mem*_ Once *U* reaches a threshold value *thr*, that we set equal to 0.5, the neuron emits a spike, *S*_*outt*_ = 1, and *U* is reset. The equations expressing the behavior of the network can be summarized by:

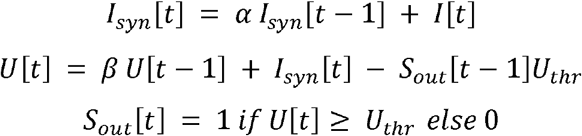

Where *I*_*syn*_ represents the synaptic current, *I* is the input current, *U* is the membrane potential of the neuron, *U* _*thr*_ the threshold value of the membrane potential and *S* _*out*_ a binary value representing the output at the indicated time instant. The last equation indicated the spike generation mechanism, while the term *S*_*out*_ [*t*−1]*U*_*thr*_ in the second equation represents the reset mechanism.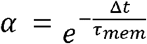 and 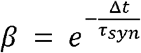 reflect, respectively, the synaptic current and membrane potential decay rates, whose biological shape is simulated using exponential functions. Both are defined as function of the respective time constant. We fixed the value of α and β respectively equal to 0.95 and 0.98.

The network is simulated using a time step of 2.5 ms, after binning the original spike arrays as describe in the spike encoding section of the methods. For what concerns the training of the network, to address the non-differentiability associated with the Heaviside function, which represents the spike generation mechanism, synaptic weights and neuron threshold are trained using the Heaviside function as surrogate gradient in the backward step^64^. We trained the network using mean square error spike count loss function and Adam optimizer, with learning rate *lr*=10^-3^. This loss encourages the target neuron to fire at a specific target frequency, while simultaneously inhibiting the firing rates of all the other output neurons to another value. The target frequency is a critical hyperparameter that requires adjustment based on the selected input time window width. Since we used data samples of 100 ms (downsampled at 750 Hz), we used a target firing ration of 0.8, which means that we expect the correct output neuron to fire at each time step with a probability of 80%. Likewise, we set the incorrect firing rate for non-target output neurons to 0.001, to reduce excessive modifications in the weights. As a performance measure we used the accuracy rate function in SNNTorch, which first predicts as the correct label the one associated to the output neuron with the higher firing rate, and then compares it with the target after being converted into a one-hot-encoding. The network was trained on 500 epochs, using a batch size of 70 data samples.

### Sensory Maps

Sensory maps were constructed based on the projected fields maps evoked via intraneural stimulation for each participant. The same 56 electrode channels used for recording were also accessible for stimulation. The electrodes were connected to an external multichannel controllable neurostimulator, the STIMEP (Axonic, and University of Montpellier). The scope of this procedure was to determine the location of the electrically-evoked sensation, as described by Petrini et al^65^. When the participant perceived any electrically evoked sensation at the minimum charge (i.e., perceptual threshold), the location was reported by the participants. This was repeated five times per channel and then averaged. All the data were collected using a custom-designed psychometric platform for neuroprosthetic applications, which allowed us to collect data using a standardized procedure^66^. These maps were used to calculate the number of active sites evoking sensations in the knee/calf versus ankle/heel versus foot/toes areas for both S1 and S2.

### Statistical Analysis

All data were exported and processed offline in MATLAB (R2024a, The MathWorks, Inc., USA) and reported as mean value ± SD (unless elsewise indicated). All statistics were performed using the available built-in functions. Since all the indicated decoders performance (SVM, MLP and SNN) are the mean performance on 5-Fold cross validation using the same data split, we compared the achieved test accuracies using pairwise t-test. The normality of data distributions was verified. In case of Gaussian distribution, two-tailed analysis of variance (ANOVA) test was applied. Elsewise, we performed the Kruskal Wallis test. Fisher’s exact test (P) was used to compare the percentages. Post hoc correction was executed in case of multiple groups of data. Additional details about the number of repetitions (n), and p-values for each experiment are reported in the results and in the corresponding figure legends.

## Data Availability

All data produced in the present study are available upon reasonable request to the authors.

## Acknowledgments

The authors are deeply grateful to the three subjects who freely donated months of their life for the advancement of knowledge and for a better future for leg amputees. The funder had no role in the experimental design, analysis, or manuscript preparation or submission. All authors had complete access to data. All authors authorized submission of the manuscript, but the final submission decision was made by the corresponding authors. This project has received funding from the European Research Council (ERC) under the European Union’s Horizon 2020 research and innovation program (FeelAgain grant agreement no. 759998).

## Author contributions

C.R. analyzed the neural data, implemented and tested the SNN-based motor decoders, prepared the figures and wrote the paper; T.S. and P.C. developed the TIME and delivered technical assistance for the human implantation and explanation procedures and reviewed the manuscript; M.B. performed the human surgeries and was responsible for all the clinical aspects of the human study; S.R. designed the study, performed and supervised the human experiments, wrote the paper; E.D., supervised all the analyses, and wrote the paper; G.V. developed the recording software for human experiments, performed the human experiments, supervised all the analyses, prepared the figures and wrote the paper. All authors edited and proofread the manuscript.

## Data and materials availability

The authors declare that the data supporting the findings of this study are available within the article and its supplementary information files. Software routines developed for the analysis are available from the corresponding author. Other data can be made available to qualified individuals for collaboration provided that a written agreement is signed in advance between the included consortium and the requester’s affiliated institution.

## SUPPLEMENTARY MATERIALS

**Fig. S1.**
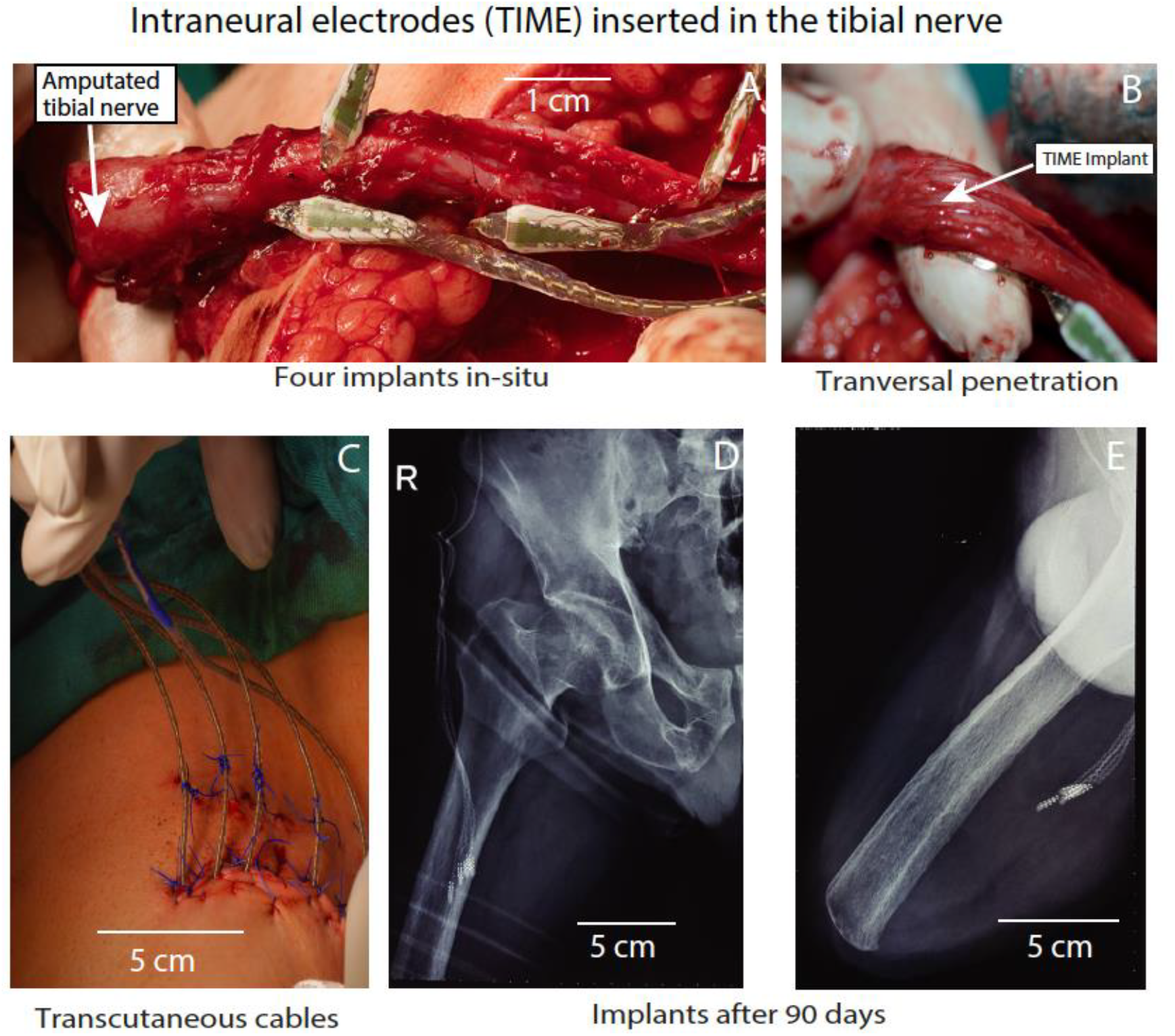
Surgical implantation of the intraneural electrodes in the tibial nerve. A-B) The electrodes are positioned to cross tranverally the amputated tibial branch of the sciatic nerve. C) The electrode cables are tunneled through the thigh and pulled out of the leg through small incisions just a few centimeters below the iliac crest, to enable transcutaneous connection with the neurorecorder. D-E) The placement of the implants within the thigh is shown in the X-ray pictures. Adapted from Petrini et al.^21^.

**Fig. S2.**
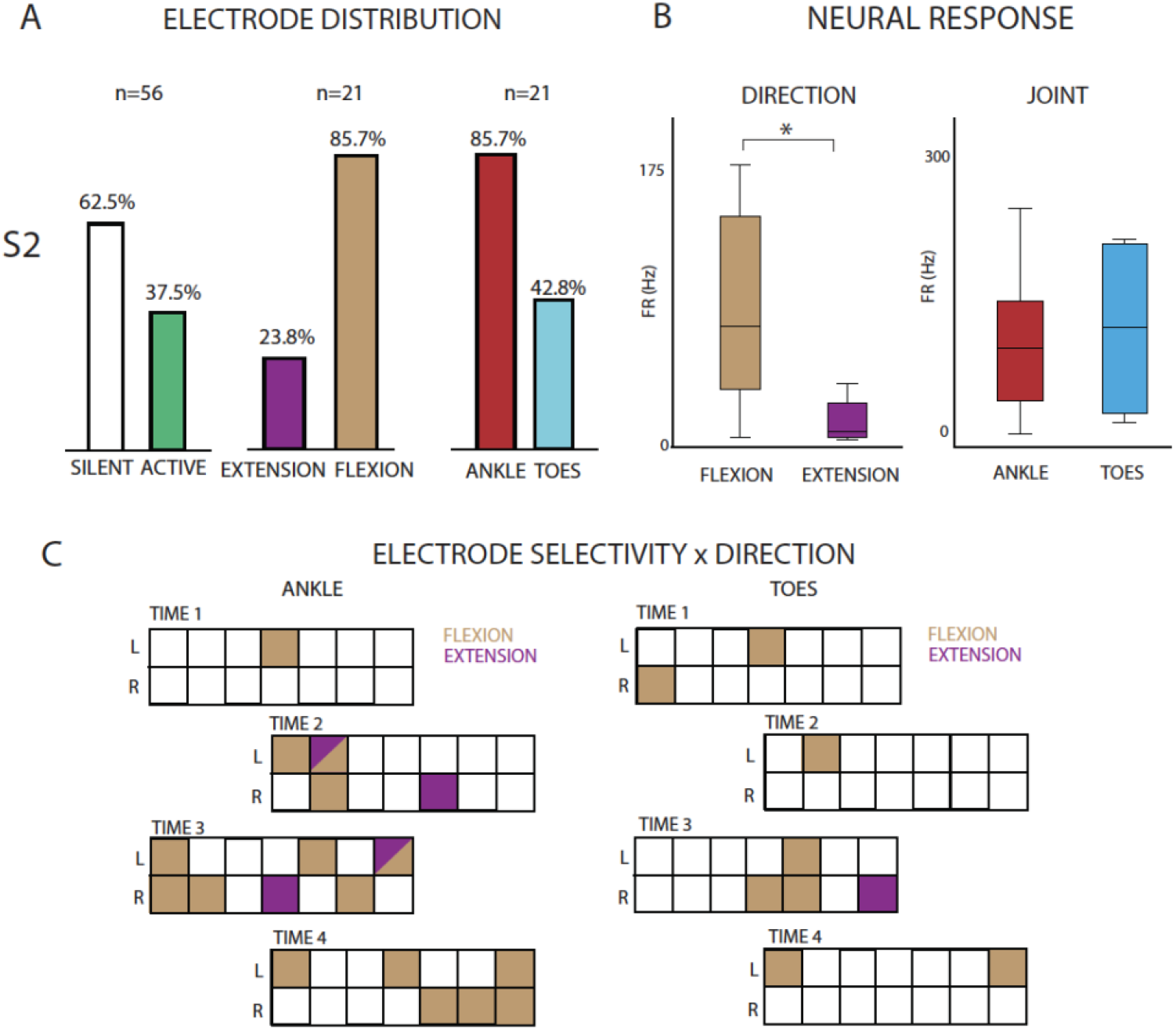
Neural response during phantom movements in S2. A) Percentage of significant modulation among all 56 channels. Data for silent vs active, flexion vs extension and knee vs ankle vs toes are reported. n indicates the sample size. B) Average firing rates across all channels for each direction and joint involved. On each box, the central mark indicates the median, and the bars the interquartile ranges, IQR. Points indicate the outliers. C) Each of the 4 TIME are reported for the 3 joint movements, showing the significant modulation for each individual channel. A channel colored in purple shows only significant modulation for flexion, in brown only for extension, both colors both directions and white no modulation. ^*^p<0.05. Data from S2.

**Fig. S3.**
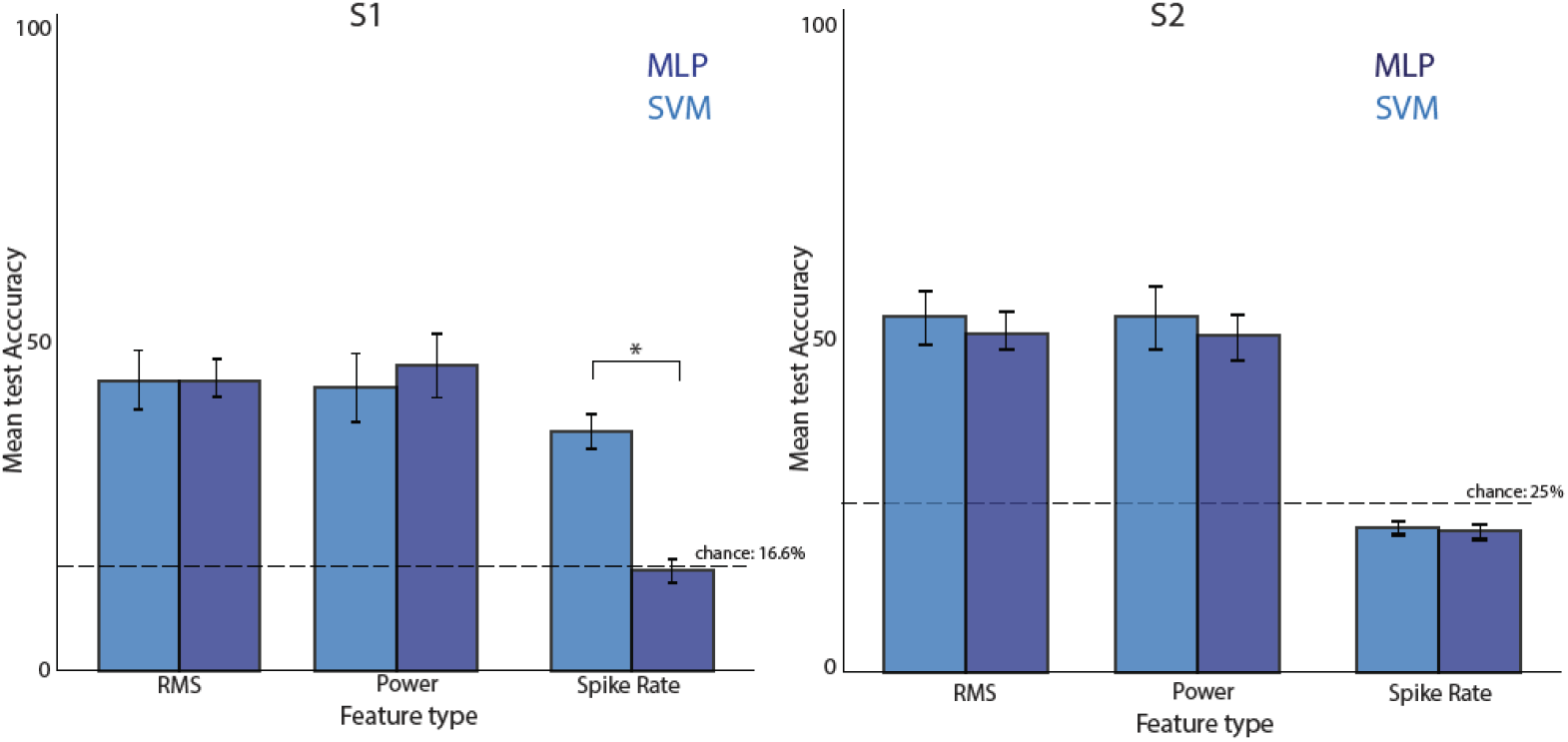
Performance of conventional neural decoders using different signal features. Comparison of accuracy of MLP and SVM decoders across various feature types commonly extracted from neural signals, including time-domain (e.g., RMS), frequency-domain (e.g., power spectral density), and spiking-based features. Results of S1 and S2 highlight how feature selection impacts the overall performance of both decoders, with some features yielding significantly higher accuracy than others across subjects. Error bars represent standard deviation across trials. Chance levels are reported with dashed lines for both participants (S1: 16.6%; S2: 25%).

**Fig. S4.**
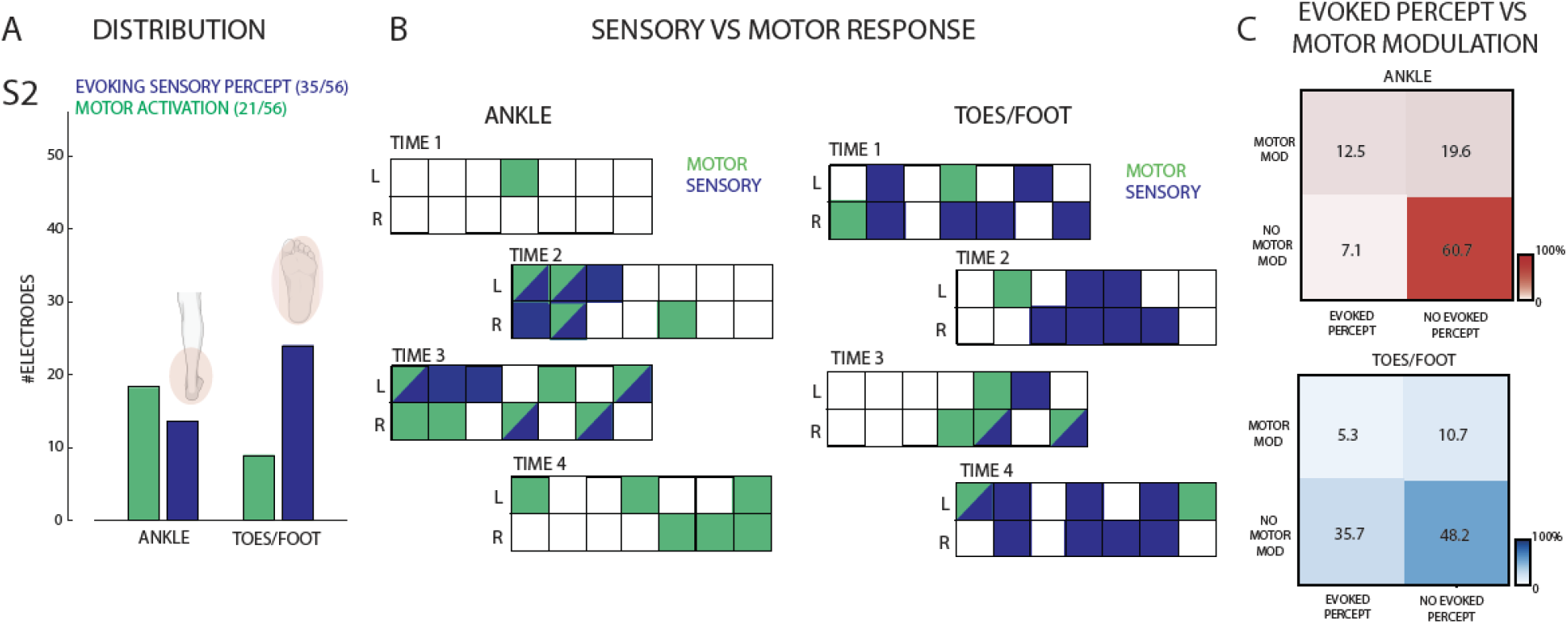
Motor modulation and sensory restoration in S2. A) Distribution of the electrodes showing significant motor modulation compared to those evoking sensation in the ankle or foot areas. B) Each of the 4 TIME are reported for the 2 joints, showing the significant motor modulation (blue) or sensory evoked response (green) for each individual channel. A channel colored in blue shows only significant motor modulation, in green only for evoked-sensation, both colors both responses and white no response. C) Modulation matrices showing the percentage of electrodes significantly modulating only during movement, evoking only sensations or both for the two joints. N=56. Data from S2.

**Table S1.** Participants’ demographics.

| SUBJECT | CAUSE OF AMPUTATION | LEVEL AND SIDE OF AMPUTATION | AGE | AMPUTATION TIME | GENDER |
| --- | --- | --- | --- | --- | --- |
| S1 | trauma | distal two-thirds of the right thigh | in his 40's | 2 | male |
| S2 | trauma | distal two-thirds of the left thigh | in his 50's | 7 | male |

**Table S2.**
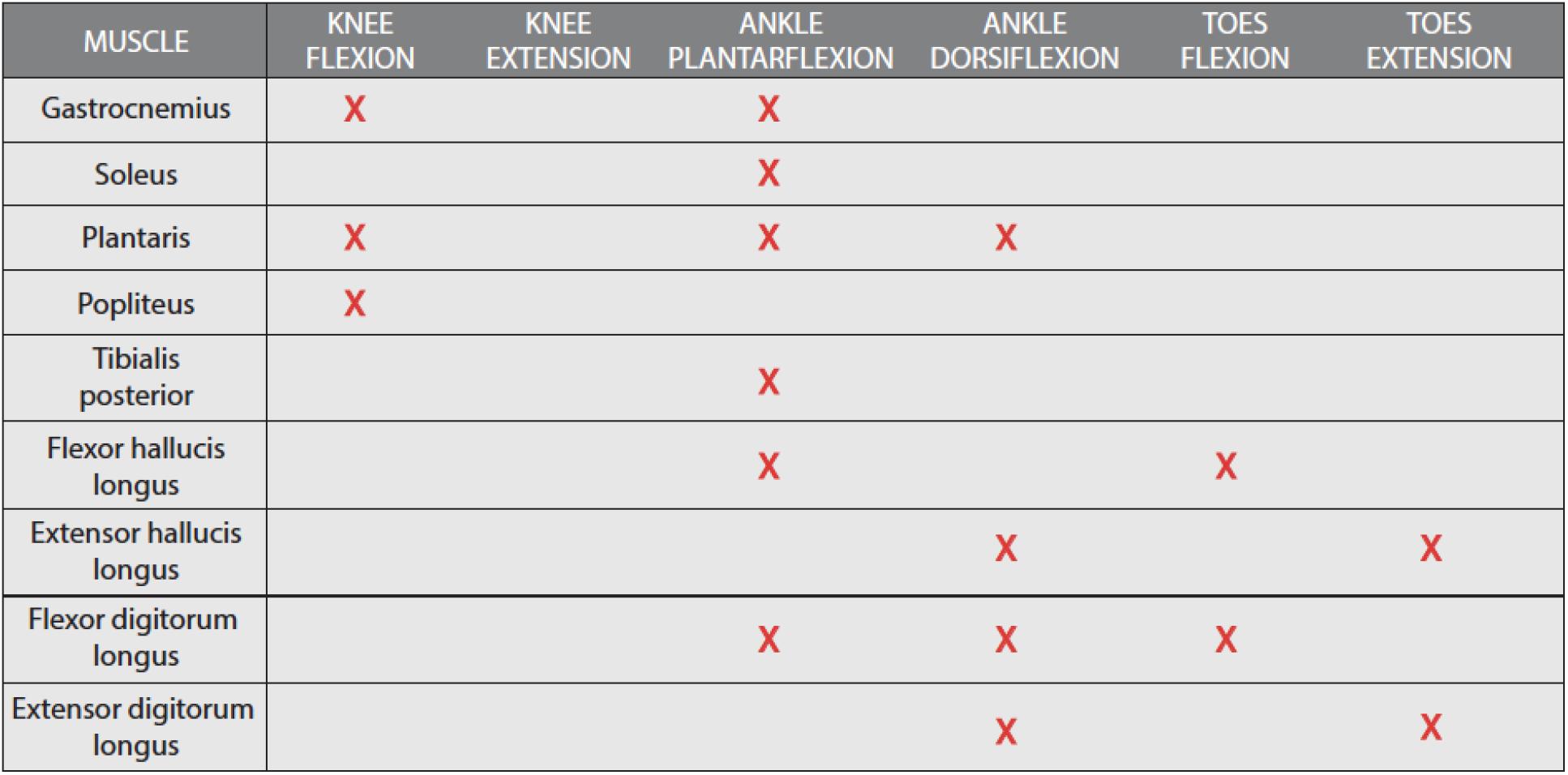
Muscular innervation of the tibial branch of the sciatic nerve.

| MUSCLE | KNEE FLEXION | KNEE EXTENSION | ANKLE PLANTARFLEXION | ANKLE DORSIFLEXION | TOES FLEXION | TOES EXTENSION |
| --- | --- | --- | --- | --- | --- | --- |
| Gastrocnemius | X |  | X |  |  |  |
| Soleus |  |  | X |  |  |  |
| Plantaris | X |  | X | X |  |  |
| Popliteus | X |  |  |  |  |  |
| Tibialis posterior |  |  | X |  |  |  |
| Flexor hallucis longus |  |  | X |  | X |  |
| Extensor hallucis longus |  |  |  | X |  | X |
| Flexor digitorum longus |  |  | X | X | X |  |
| Extensor digitorum longus |  |  |  | X |  | X |

